# Rate and synchrony at the level of the auditory nerve are not sufficient to account for behaviorally estimated cochlear compression - a modeling study

**DOI:** 10.1101/2020.04.06.20055855

**Authors:** Gerard Encina-Llamas, Jens Thuren Lindahl, Bastian Epp

## Abstract

Methods based on psychoacoustical forward masking have been proposed to estimate the local compressive growth of the basilar membrane (BM). This results from normal outer hair cells function, which leads to level-dependent amplification of BM vibration. Psychoacoustical methods assume that cochlear processing can be isolated from the response of the overall system, that sensitivity is dominated by the tonotopic location of the probe and that the effect of forward masking is different for on- and off-characteristic frequency (CF) maskers. In the present study, a computational model of the auditory nerve (AN) in combination with signal detection theory was used to test these assumptions. The underlying idea was that, for the BM compression to be estimated using psychoacoustics, enough information should be preserved at the level of the AN, because this forms an information bottleneck in the ascending auditory pathway. The simulated AN responses were quantified in terms of rate and synchrony for different types of AN fibers and CFs. The results show that, when using a low-intensity probe, local activity at the tonotopic location of the probe frequency is the dominant contributor to sensitivity in the healthy auditory system. However, on- and off-CF maskers produced similar forward masking onto the probe, which was mainly encoded by high- and to little extent by medium-spontaneous rate fibers. The simulation results suggested that the estimate of compression based on the behavioral experiments cannot be derived from sensitivity at the level of the AN but may require additional contributions, supporting previous physiological studies.

## Introduction

The healthy mammalian auditory system is remarkable in its ability to handle a large dynamic range of incoming sound levels while preserving high sensitivity to low-intensity sounds. One contributor to this ability is the nonlinear processing at the level of the basilar membrane (BM). Measurements in non-human animal models have demonstrated that the BM movement peaks at a specific location when stimulated with a pure tone of a given frequency and level. This frequency is referred to as the “characteristic frequency (CF)” of that location. This results in a tonotopical organization of CFs along the BM, which links each BM location to one frequency (Bekesy 1947). The assignment of a CF to one location is usually done for a pure tone of low and fixed level, even though the peak of the movement shifts towards the base as level increases (Ruggero et al. 1997). At or near the CF of a pure tone stimulus (on-CF), the BM velocity at the corresponding location grows compressively with increasing sound level (for a review, see Robles and Ruggero 2001). At locations corresponding to CFs more than 1/2-octave below or more than 1/3-octave above that of the stimulus frequency (off-CF), the BM velocity (in chinchillas) was shown to grow linearly with increasing sound level (Figs. 6 and 7 in Ruggero et al. 1997). This on-CF compressive growth is commonly assumed to be the result of an “active mechanism”, presumably due to the electromotility of the outer hair cellss (OHCs) in the cochlea (e.g., Gummer et al. 2002; Dallos 2008; Ashmore 2008; Dong and Olson2013; Guinan 2013). OHCs induce on-CF level-dependent gain to the BM motion leading to the high sensitivity to low level stimuli. Such a “cochlear amplifier” was proposed to reduce its gain as stimulus level increases, which would explain the on-CF compressive input/output (I/O) function. This level-dependent amplification would also effectively increase the sensitivity of the mechano-electrical transduction mechanism in the inner hair cellss (IHCs) (e.g., Dallos and Harris 1978; Guinan 2011).

**Figure 1.**
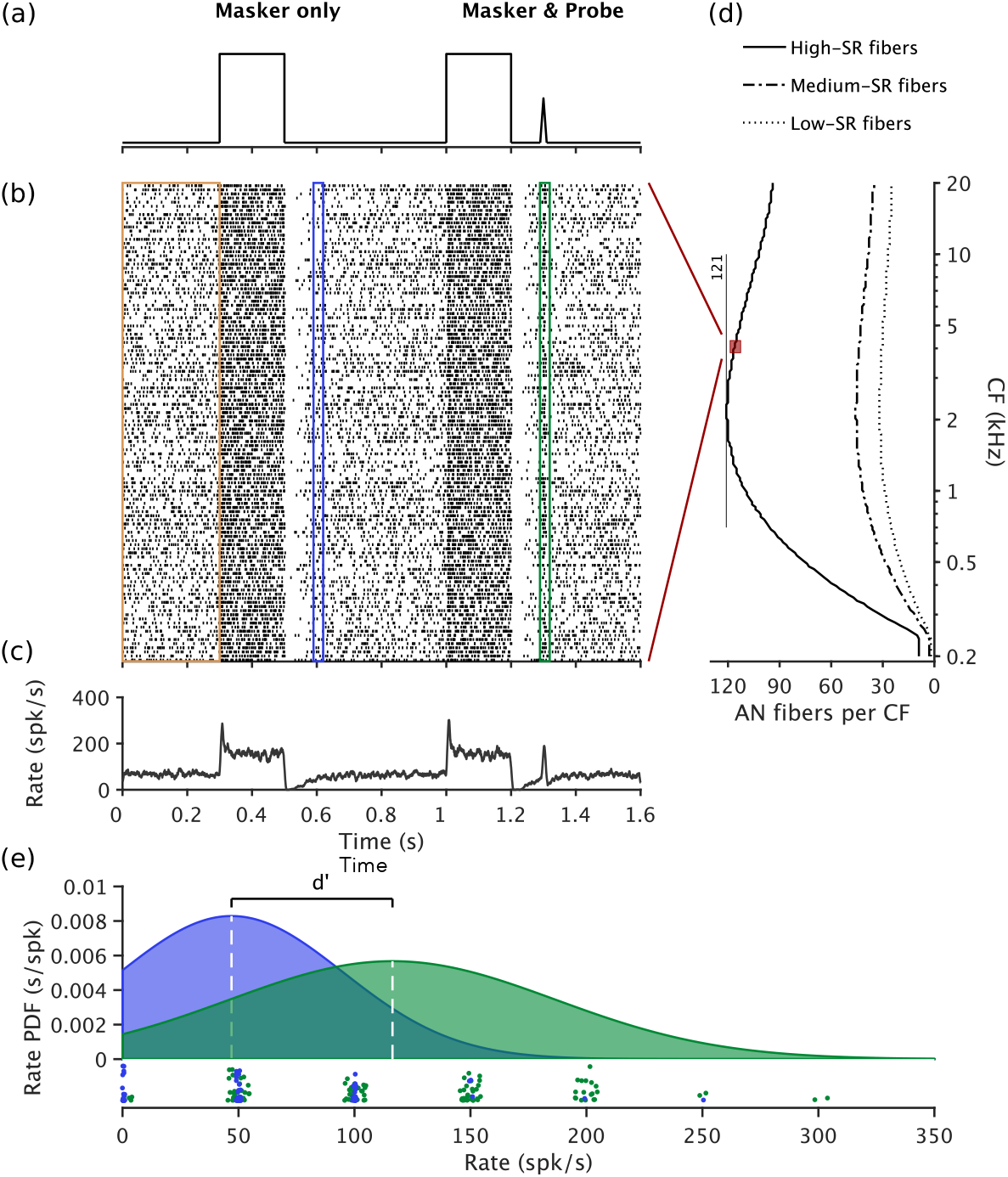
(a) Sketch of the stimuli. Stimuli start with 300 ms without sound (quiet) from which SR can be computed (yellow vertical rectangle in (b)). The masker-only block was presented with 500 ms of post-masker silence to allow the neurons to recover. Then the same masker was presented followed by the probe with varying MPI (from 2 to 100 ms; 90 ms in the present example). Maskers were presented at the probe frequency (on-CF, 4 kHz) or at 0.6 times the probe frequency (off-CF, 2.4 kHz), with masker level ranging from 10 to 110 dB SPL (30 dB SPL in the present example). (b) Raster plot of simulated AN neurons for the CF of 4 kHz. The vertical rectangles indicate the analysis windows for the estimation of SR (yellow), the probe response window when no probe was present (blue) and the probe response window with the probe present (green). (c) Resulting summed neuronal response from all the simulated AN fibers at this CF (comparable to a peri-stimulus time histogram (PSTH)). The simulations show clear AN properties such as spontaneous activity, onset response (short-term adaptation) and the reduction in rate post-masker (long-term adaptation), as well as the response to the probe. (d) Distribution and number of simulated AN fibers across CF for each AN fiber type. (e) Fitted probability density function (PDF) of the rates computed at each probe analysis window (with the probe present and absent) and individual data points shown as a rugplot. White vertical dashed lines indicate the mean of each fitted distribution that is used in the *d*′ calculation.

**Figure 2.**
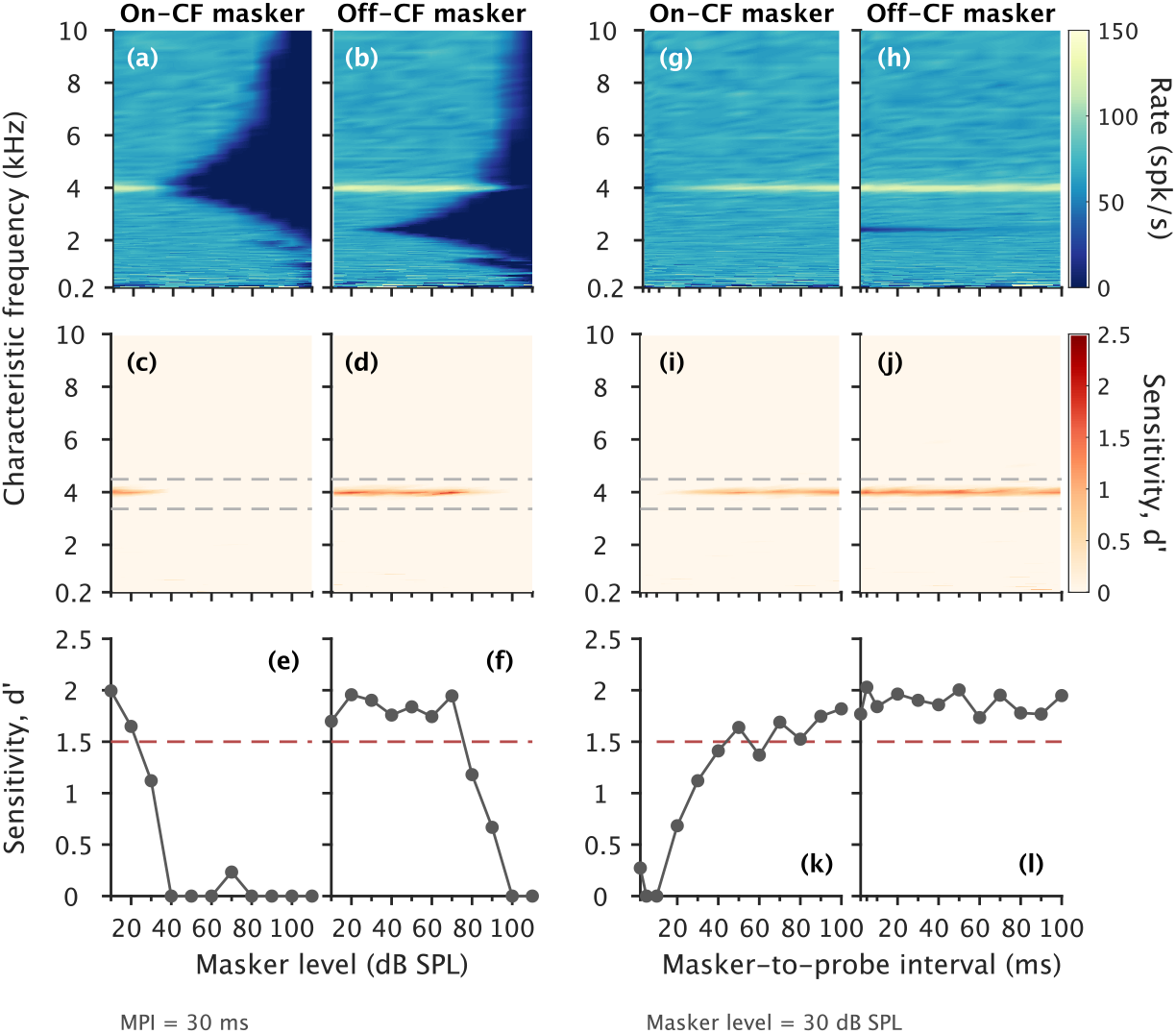
Exemplary simulation results for different masker levels at a MPI of 30 ms (panels (a-f)), and as a function of MPI for a masker level of 30 dB SPL (panels (g-h)), for the case of NH. Results show simulations considering only rate and HSR fibers. Panels (a, b) show heatmaps of rate as a function of CF and masker level, for on-CF and off-CF maskers, respectively. Higher rates due to the probe were concentrated near on-CFs. Rate decreased with increasing masker level at different ranges of masker levels for the on-CF and off-CF maskers. Panels (c, d) show *d*′ as a function of CF and masker level, for on-CF and off-CF maskers, respectively. Grey-dashed horizontal lines indicate the on-CF range. Panels (e, f) show combined *d*′ as a function of masker level, for on-CF and off-CF maskers, respectively. Red-dashed horizontal lines indicate the *d*′ threshold 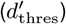. Panels (g, h) show rate as a function of CF and MPI using a masker of 30 dB SPL, for on-CF and off-CF maskers, respectively. Rate increased with increasing MPI for the on-CF masker. Similar progression was observed for *d*′ as a function of CF and MPI (panel (i)) and for the combined *d*′ as a function of MPI. For the off-CF masker, rate was constantly high at all MPI (panel (h)), as well as *d*′ (panel (j)) and combined *d*′ (panel (l)).

**Figure 3.**
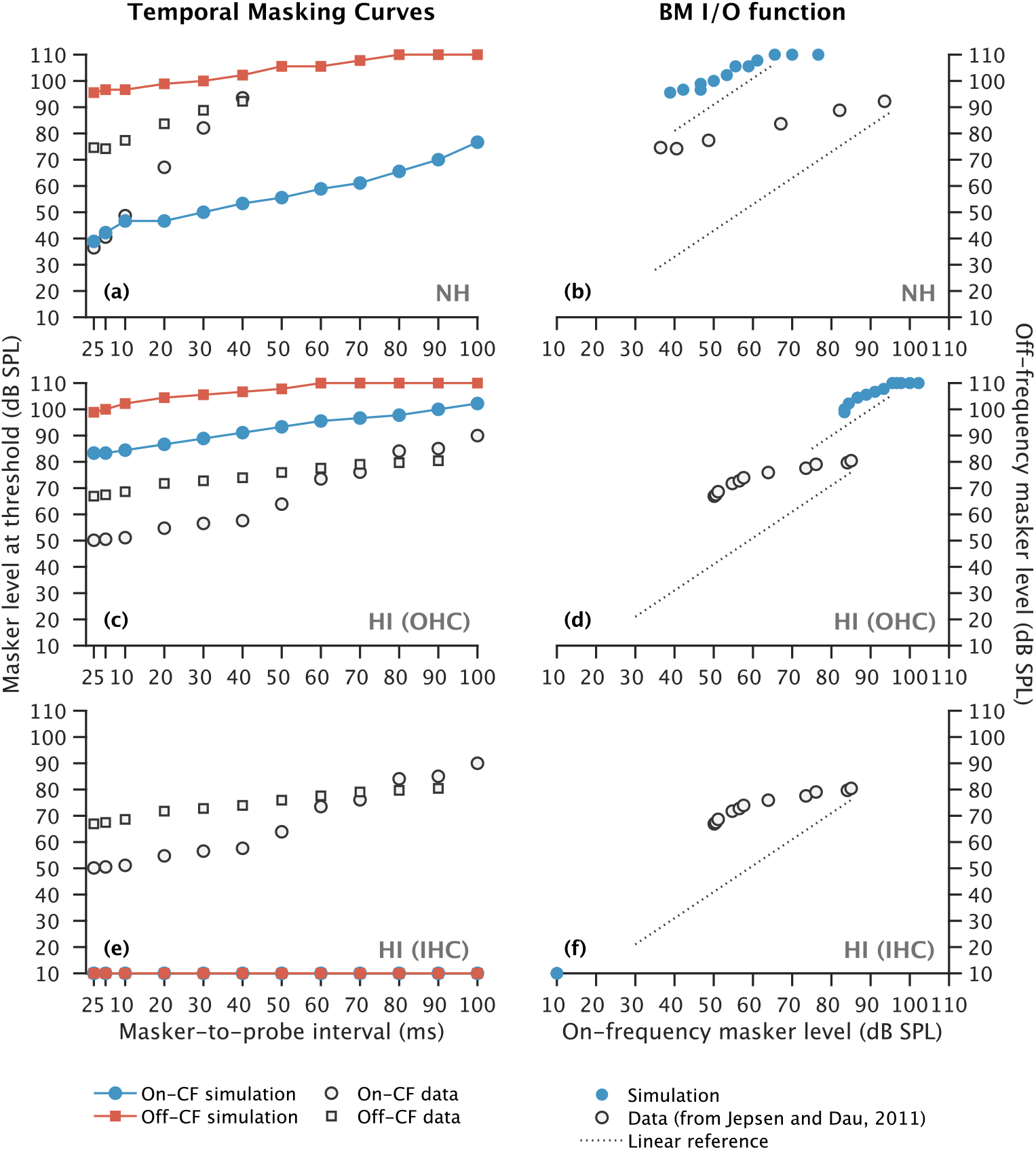
Simulated TMCs (leftmost panels) and corresponding BM I/O functions (rightmost panels) for the NH listener (panels (a, b)), for the HI listener assuming only OHC dysfunction (panels (c, d)), and for the HI listener assuming only IHC dysfunction (panels (e, f)). Blue circles in the leftmost panels show on-CF TMCs and red squares show off-CF TMCs. The 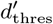 were 1.5 and 3 for the NH and the HI (only OHC) listeners, respectively. Open circles and open squares show behavioral results from Jepsen and Dau (2011) (reprinted with permission from the authors and the publisher). Blue circles in the rightmost panels show the corresponding simulated BM I/O functions resulting from plotting the off-CF TMC as a function of the on-CF TMC. Open circles show the corresponding behavioral BM I/O functions.

**Figure 4.**
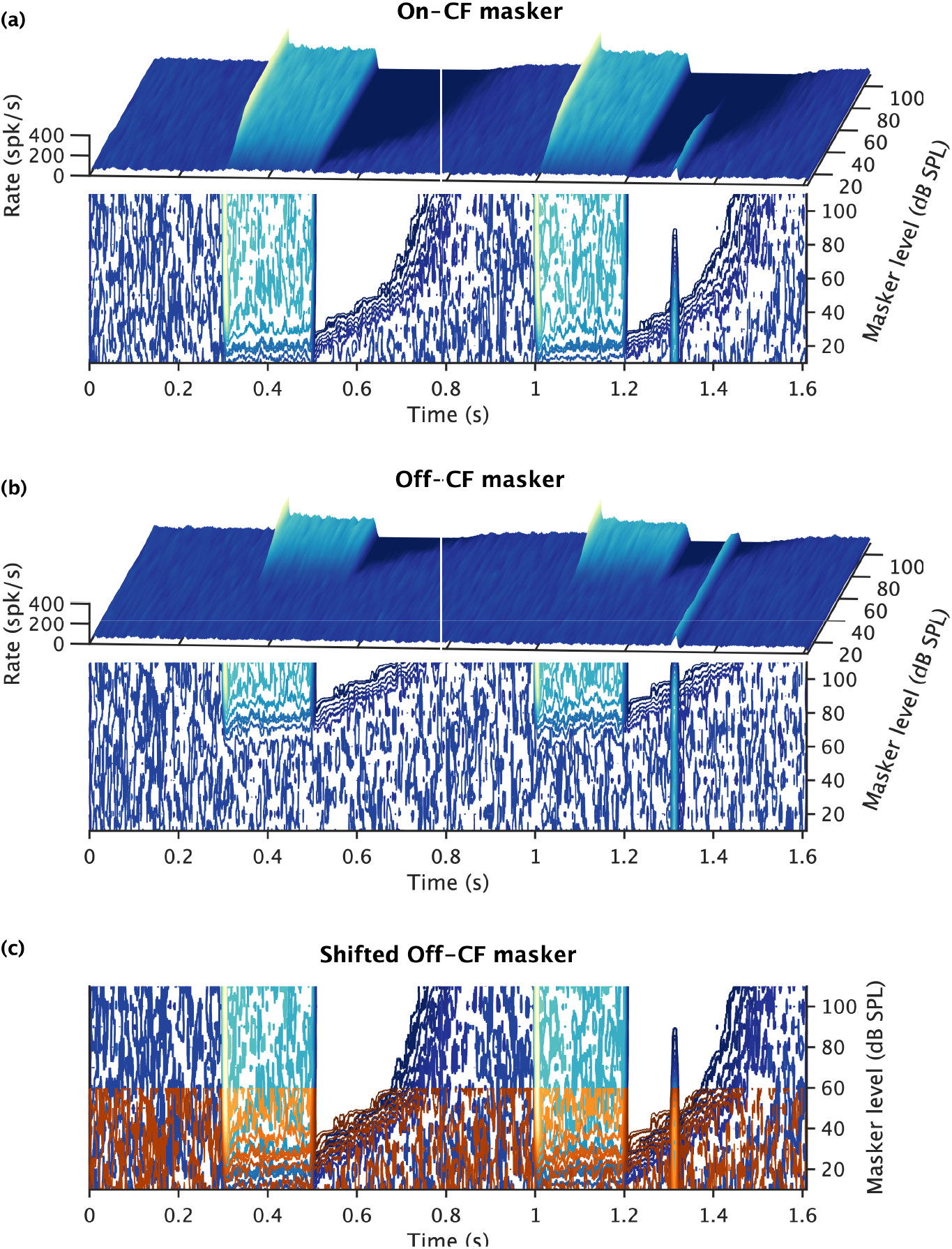
Rate post-masker recovery time course analysis for the HSR AN fibers tuned at the location tuned to on-CF. Panel (a) and (b) show simulated AN rate evoked by the whole stimuli as a function of time (abscissa) and masker level (ordinate) for on- and off-CF maskers, respectively. The results are shown as a surface plot as well as contour plot with iso-rate slices. In panel (a), the reduction in rate due to the masker can be seen as a dark-blue shadow at the masker offset. Such reduction resulted in a reduction of the activity evoked by the probe after the second masker (at time about 1.3 s). In panel (b), the activity evoked by the maskers was visible at much higher masker levels, resulting in a no reduction of the activity evoked by the probe. Panel (c) shows the same contour plots for the on- and off-CF maskers but shifting the off-CF masker response by 60 dB along the masker level axis.

**Figure 5.**
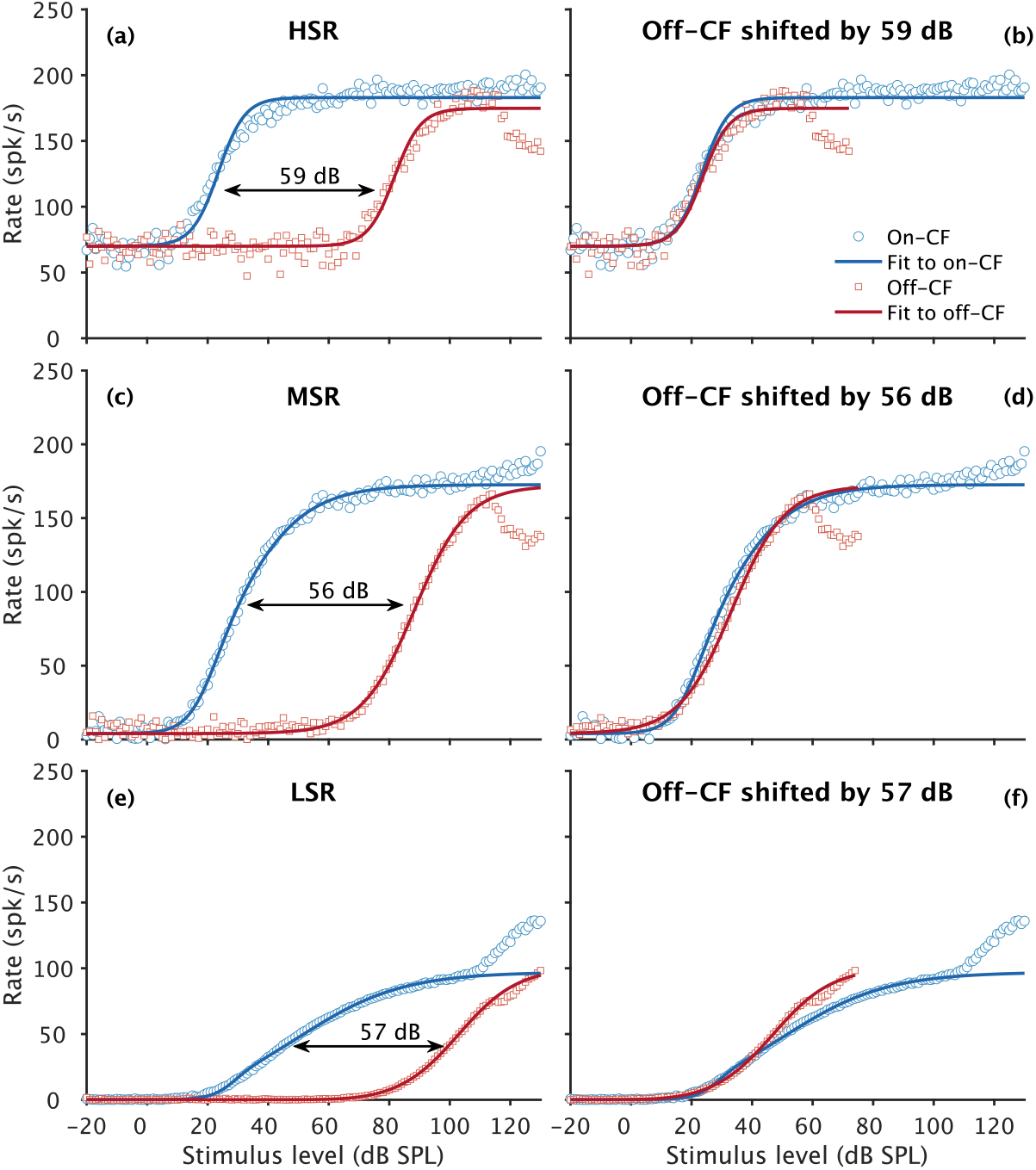
Simulated rate-level functions of AN neurons tuned at on-CF for the on-CF masker (blue circles) and the off-CF masker (red squares). Panels (a, b), (c, d) and (e, f) show rate-level function for HSR, MSR and LSR, respectively. The leftmost panels ((a, c, e) show the original rate-level functions whereas the rightmost panels ((b, d, f)) show the same rate-level functions but with the off-CF masker rate-level function shifted in the stimulus level axis so that on- and off-CF rate-level functions overlap at a rate of 30 spk/s above threshold. Solid lines indicate fits to the rate-level functions up to a stimulus level of 100 dB SPL, as described in Yates et al. (1990),

In non-human animal models, OHCs were shown to be vulnerable to noise exposure (Liberman and Kiang 1978). Damage of OHCs leads to an impairment of the active mechanism and reduces sensitivity at on-CF, resulting in a loss of compression (see the elevated thresholds only at the tip of the auditory nerve (AN) tuning curves in the right panels of Fig. 14 in Liberman and Dodds 1984). Behaviorally, OHC deficits in human listeners have been associated with sensorineural hearing impairment, typically characterized by elevated threshold in quiet (i.e., a decrease in sensitivity), poorer frequency selectivity and abnormal loudness growth with sound pressure level (SPL) (Plack 2013). Because of the connection between OHC health and BM compression, it was argued that behaviorally estimated compression can be a proxy for the state of the OHCs and therefore for the health of the inner ear (e.g., Oxenham and Bacon 2003).

Diagnosis of hearing impairment and fitting of hearing aids are usually based on psychoacoustical estimates of hearing threshold through pure-tone audiometry. The basic idea behind amplification through a hearing aid is to compensate for the gain loss reflected by the increased hearing threshold. Hence, the implicit assumption is that OHC damage is the dominant source of hearing impairment. The fact that not all hearing impaired (HI) listeners benefit equally much from wearing hearing aids indicates that other elements than OHC health might be relevant. It has been reported in non-human animal models that pure-tone audiometry is highly insensitive to large losses of IHCs (Lobarinas et al. 2013), and to large losses of the synaptic connections between the IHCs and the AN fibers (i.e., cochlear synaptopathy, Kujawa and Liberman 2009). Furthermore, a recent histopathological study in human temporal bones suggested that audiometric thresholds and hair cell damage (both IHCs and OHCs) are not significantly correlated (Landegger et al. 2016). Newer imaging techniques have suggested, however, that hair cell damage may has been underestimated and that a reanalysis of the data could result in a higher correlation between hair cell damage, specially OHC damage, and audiometric thresholds (Wu et al. 2020). Experimental audiology is aiming towards precise and individualized diagnostics (Lopez-Poveda et al. 2017) in order to provide individualized compensatory strategies (Johansen et al. 2018). To fulfill this goal, methods that provide accurate estimates of IHC and OHC status are needed. In living humans, invasive assessment of hair cell status is not possible due to ethical constraints. To overcome this limitation, estimates of cochlear compression, based on psychoacoustical forward masking paradigms, have been proposed as a potential diagnosis tool to identify and differentiate OHC and IHC damage (Oxenham and Bacon 2003; Lopez-Poveda and Johannesen 2012; Johannesen et al. 2014).

Two methods proposed to estimate compression are the growth of masking (GoM) and the temporal masking curve (TMC) paradigms (Oxenham and Plack 1997; Nelson et al. 2001; Oxenham and Bacon 2003; Plack 2013). Both methods use a combination of on- and off-CF tonal maskers in connection with a brief tonal probe. To overcome the problem of spread of excitation produced by the probe, TMCs were proposed as an alternative to GoM (Nelson et al. 2001). In TMCs, the probe level is kept at a low sound level near hearing threshold while the masker level is varied to find masking threshold for varying masker-to-probe intervals (MPIs). Both methods assume that the excitation of the BM grows compressively with masker level for an on-CF masker, and linearly with masker level for an off-CF masker. It is assumed that, for an off-CF masker, the level required to mask the probe at different MPIs is only dependent on the recovery from forward masking because the growth of the masker is linear, while for an on-CF masker, masker level at threshold depends on the recovery of forward masking (assumed to be the same for on- and off-CF) plus the compressive growth of the on-CF masker. Several studies have reported good correspondence between behaviorally estimated cochlear compression and the slope of BM velocity-intensity functions directly recorded in non-human mammals (e.g., Nelson et al. 2001; Lopez-Poveda et al. 2003; Plack et al. 2004; Lopez-Poveda et al. 2005; Lopez-Poveda and Alves-Pinto 2008; Jepsen and Dau 2011; Lopez-Poveda and Johannesen 2012; Johannesen et al. 2014; Fereczkowski et al. 2017).

It has been argued that accurate models of the auditory periphery and quantitative simulations could assist in predicting the mechanisms underlying the perceptual measures (Oxenham and Bacon 2003). Hybrid models containing physiological parts coupled with psychoacoustical concepts (e.g., the temporal integration window model by Oxenham and Moore 1994) were shown to be successful in accounting for the measured TMCs, both in normal hearing (NH) and HI listeners, used to estimate BM compression (Jennings et al. 2014). However, physiological studies have demonstrated that behavioral forward masking does not fully correspond to forward masking observed at the level of the AN (Relkin and Turner 1988), and that it agrees better with forward masking observed in neurons at the level of the inferior colliculus (IC) (Nelson et al. 2009).

In the present study, a state-of-the-art computational model of the AN (Bruce et al. 2018) in combination with methods from signal detection theory (SDT) was used to study the potential physiological underlying mechanisms of TMCs in the auditory peripheral system. It was hypothesized that if cochlear compression can be derived from TMCs using on- and off-CF maskers, enough information must be present already at the level of the AN, as it is the first mandatory neuronal stage post-BM and acts as an information bottleneck. The following assumptions were made within the modeling framework: 1) Perceptual outcome measures are formed using exclusively information present at the level of the AN in the form of a) spike rate and b) synchrony. 2) Perceptual outcome measures are based on temporal information encoded in any responsive AN neuron at any CF, rather than only at the on-CF of the probe tone. 3) All three types of AN fibers (high-SR (HSR), medium-SR (MSR) and low-SR (LSR)) potentially contribute to the behavioral outcome measure.

## Methods

The experimental paradigm used to simulate the response of the AN was the same as in Jepsen and Dau (2011). The AN model by Bruce et al. (2018) was used with parameters as described in Encina-Llamas et al. (2019).

### Experimental paradigm

The stimulus consisted of a tonal masker with a duration of 200 ms (gated with 5 ms rise/fall cosine ramps), a MPI (between the masker offset and the probe onset) of 2, 5 and 10 to 100 ms in steps of 10 ms, and a probe tone with a duration of 20 ms (gated with 10 ms rise/fall Hann window ramps with no steady-state portion). The frequency of the probe was 4 kHz. The masker was presented at levels from 10 to 110 dB SPL in steps of 10 dB. The probe level was fixed at 10 dB sensation level (SL) relative to the simulated model threshold. Thresholds in the AN model were calculated for each CF by performing a non-parametric one-tailed two-sample permutation test for equality of the means with 10 000 permutations. The statistical test evaluates, with a significance level of *α* = 1%, whether the distribution of spike rates obtained from 100 computations of the AN model stimulated with a pure tone at a given fiber CF is statistically higher than a distribution of spontaneous rate (SR) obtained from 100 computations with no stimulus present. The masker was either presented at the probe frequency (on-CF, 4 kHz) or at a frequency 0.6 times the probe frequency (off-CF, 2.4 kHz). First a block with the masker only (masker-only block) was presented to the model followed by a block with the same masker plus the probe (see Fig. 1(a)). The masker-only block was presented after 300 ms of simulation in quiet from which SR was estimated. After the masker-only block, no stimuli were presented for a period of 500 ms to allow the neurons to fully recover from the masker excitation (post-masker recovery time).

Two listeners from Jepsen and Dau (2011) were simulated: a NH listener represented by the averaged NH audiogram, and one mildly HI listener (HI01) with hearing threshold of 45, 50 and 45 dB hearing level (HL) at 2, 3 and 4 kHz, respectively. A combination of 2/3 of OHC dysfunction and 1/3 of IHC dysfunction was assumed in the model simulations, as suggested by Spongr et al. (1997); Plack et al. (2004); Lopez-Poveda and Johannesen (2012).

### Model of the auditory nerve

The AN model by Bruce et al. (2018) was used to simulate TMCs at the level of the AN. In short, the AN model filters the input stimulus waveform (in pascals) by a linear filter representing the middle-ear transfer function. The BM processing is modeled as a filter-bank that includes three parallel filter paths at each CF to account for the time-varying level-dependent nonlinearities due to gain induced by OHC motion. The IHC block models the deterministic IHC transmembrane potential through a rectifying non-linearity together with a low-pass filter to account for the limitation in phase-locking. The synapse model is implemented using fast and slow power-law functions to account for the offset adaptation and long-term adaptation observed in AN single unit recordings (Zilany et al. 2009). Onset (short-term) adaptation is modeled as an adaptive redocking mechanism with 4 sites of synaptic release.

The AN response was simulated for neurons with CFs ranging from 0.2 to 20 kHz, lumped into 200 CFs equally spaced along a logarithmic axis representing a discretization of the cochlea into 200 segments. The transmembrane potential of a single IHC was simulated per CF (i.e., per cochlear segment). The number of AN fibers at each CF followed a non-uniform distribution as used in Encina- Llamas et al. (2019), who considered a total number of 32000 AN fibers (based on physiological data from human temporal bones, Spoendlin and Schrott 1989) distributed over the 200 CFs. For each CF, 61 % of HSR fibers, 23 % of MSR fibers and 16 % of LSR fibers were considered, based on the physiology of the cat cochlea (Liberman 1978). Hair-cell impairment was implemented by fitting the listener’s audiogram using the *fitaudiogram2* MATLAB function implemented by Zilany et al. (2009). This function allows to define the proportion of threshold elevation that is attributed to either OHC or IHC dysfunction.

### Analysis metrics

At each CF and each AN fiber, the resulting spike trains were stored and used to compute two metrics: spike rate *r*_*L*_ and spike synchrony *ρ*(*f*). These metrics were computed over two different time windows in the simulation: one centered at the time of the probe response in the masker+probe block (green rectangle in Fig. 1(b)) and a second centered at the time of the probe response in the masker-only block in the absence of the probe (blue rectangle in Fig. 1(b)). Analysis was also performed considering SR extracted from the first 300 ms of the simulation. Spike rate *r*_*L*_ (equation 1) was calculated summing the number of spikes *N*spikes normalized by the length of the analysis window *L*. Spike synchrony *ρ*(*f*) (equation 2) was calculated by the vector sum of spike times projected onto a phasor rotating on the unit circle in the complex plane at the probe frequency *f*.

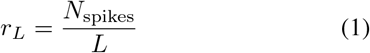

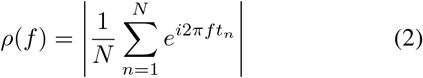

For each CF and fiber type, distributions of rate and synchrony were obtained for each window of analysis (Fig. 1(e)). The resulting distributions were used to calculate the sensitivity index *d*′ using their mean and variance values, as defined in equation (3) (Jones 2016).

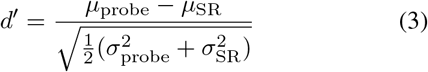

All AN fibers were treated as independent sources of information. Information in the form of *d*′ was combined across fiber type, across CFs within a 2-octave range centered at the frequency of the probe (4 kHz) and across metrics (rate and synchrony). The final combined sensitivity 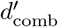 for each group of parameters was calculated as defined in equation (4), with SR = 1, 2, 3 denoting the AN fiber type (i.e., HSR, MSR and LSR fibers, respectively), CF_k_ denoting the CF index, and m = 1, 2 denoting the two derived metrics (rate and synchrony, respectively).

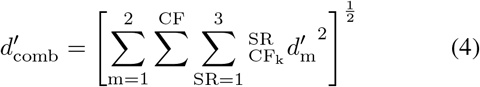

## Results

### Simulations as a function of masker level and masker-to-probe interval

Figure 2 shows exemplary simulation results for the NH listener. Results are shown for different masker levels and a constant MPI of 30 ms (panels (a-f)), and for a constant masker level of 30 dB SPL and for different MPIs (panels (g-l)). Results in Fig. 2 are only based on rate and HSR fibers. Results for MSR and LSR AN fibers can be found in Supplementary Figs. 6 and 7, respectively. The combined *d*′ in panels (e, f, k, l) was calculated across CFs within a 2-octave range centered at the frequency of the probe.

Panels (a, b) show heatmaps of rate evaluated in the analysis window of the probe response (green vertical rectangle in Fig. 1(b)) as function of CF (ordinate) and masker level (abscissa), for the on-CF and off-CF maskers, respectively. In panel (a), the low level of the probe produced a narrow-band response of high rate concentrated at around 4 kHz. This rate reduced rapidly as masker level increased. For masker levels higher than about 40 dB SPL, the rate reduced to values below SR, reaching 0 spk/s. In panel (b), the rate evoked by the probe remained high up to a masker level of about 80 dB SPL. For masker levels above 80 dB SPL, rate decreased, similar to (a). Such reduction in rate (dark-blue areas in panels (a, b)) occurred at much higher masker levels for the off-CF masker than for the on-CF masker at the on-CF location. For an on-CF masker, the location of probe excitation coincided with the tip of the rate reduction pattern produced by the masker (panel (a)). In contrast, an off-CF masker excited the AN fibers tuned at the probe frequency (on-CF) in their tails (panel (b)). Thus, masking the probe required higher intensities for an off-CF masker than for an on-CF masker. This resulted in simulated TMCs with higher masker levels at threshold for off-CF maskers than for on-CF maskers for the NH listener (Fig. 3 (a)). The level required for an on-CF masker to mask the probe was higher in the case of impairment due to OHC dysfunction than in the NH listener (panels (a) and (b) in Fig. 3). This is because OHC dysfunction leads to a broadened and elevated tip of the AN tuning curves (Liberman and Dodds 1984) (see Supplementary Fig. 8). Besides the threshold elevation at the tip of the tuning curves, the growth of rate above threshold (i.e., rate-level function) for the on-CF masker in the case of hearing impairment due to only OHC dysfunction was similar to the NH case but shifted to higher stimulus levels (see Supplementary Fig. 14). On the contrary, IHC dysfunction, not only resulted in elevated threshold, but the growth of rate above threshold was also reduced, both for the on- and the off-CF maskers (see Supplementary Fig. 15). This led to very weak masker-induced activity that was unable to mask the probe (Fig. 3(e)).

Panels (c, d) in Fig. 2 show the calculated *d*′ as a function of CF (ordinate) and masker level (abscissa) for on-CF and off-CF maskers, respectively. High *d*′ values can be observed in the same CF range and masker level range where rate was high in panels (a, b). High *d*′ were found within the on-CF range, indicated by gray-dashed horizontal lines. The combined *d*′ in panels (e, f) covaried with the rate evoked by the probe (panels (a, b)). The reduction in combined *d*′ was very similar for the on-CF and the off-CF maskers, with a steep decreasing slope for masker levels of about 20 dB SPL and 70 dB SPL, respectively. These results clearly reflect the effect of spatial spread and reduction in rate evoked by the probe with increased masker level.

Panels (g-l) show the same information as panels (a-f) but for a constant masker level and varied MPI. The rate evoked by the probe increased with increasing MPI for an on-CF masker and was constant for an off-CF masker, in this example. The increase in rate for the on-CF masker reflects the decay of the rate reduction caused by the masker offset. The fact that the off-CF masker did not affect the rate evoked by the probe shows that the masker energy did not spread to the tonotopic location of the probe. Similar to panels (c-d) and (e-f), the *d*′ as a function of CF was high in a narrow region around the CF of the probe tone. The combined *d*′ increased in line with the change of rate and *d*′ within the on-CF range. These results reflect the temporal decay of the reduction in rate after masker offset and the difference in on-CF vs off-CF masker activity at the CF of the probe.

### Simulations of AN temporal masking curves

Figure 3 shows simulated TMCs (panels (a, c, e)) and corresponding BM I/O functions (panels (b, d, f)). Panels (a, b) show the results for the NH listener, panels (c, d) for the HI listener assuming only OHC dysfunction to account for the hearing threshold elevation, and panels (e, f) for the HI listener assuming only IHC dysfunction to account for the elevation. The simulated results are based on the combination of *d*′ across types of AN fiber, across CFs and for rate and synchrony, as defined in equation (4). A *d*′ threshold 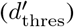 of 1.5 was arbitrarily chosen for the NH in order to detect the probe (dashed-red horizontal line in panels (e, f, k, l) in Fig. 2). Open symbols indicate the behavioral data from Jepsen and Dau (2011). The behavioral TMCs for the NH listener (panel (a)) show a steeper slope for the on-CF masker (open circles) than for the off-CF masker (open squares), leading to a derived BM I/O function (panel (b)) with a slope of about 0.3 dB/dB. The slopes of the simulated on- and off-CF TMCs were shallower than the behavioral TMCs and were more similar, which led to a simulated BM I/O function with a less compressive slope of about 0.6 dB/dB.

The behavioral TMCs for the HI listener (panel (c, e)) show shallower slopes compared to that of the NH listener (panel (a)). The TMCs show very similar slopes up to MPIs of 50 ms, where the slope of the on-CF masker increases. This led to a derived BM I/O function (panel (d, f)) with a slope close to 1 dB/dB for low masker levels and of about 0.3 dB/dB for masker levels above about 60 dB SPL. The simulated TMCs assuming only OHC dysfunction were higher in masker level at threshold compared to the behavioral data. The simulated TMCs also showed an almost parallel growth up to MPIs of 60 ms, but because the off-CF

TMC reached the maximum masker level of 110 dB SPL at longer MPIs. This led to a derived BM I/O function with a slope of about 0.7 dB/dB for levels of about 60 dB and above (panel (d)). When OHC dysfunction was applied, the range of CFs with significant *d*′ exceeded the definition of on-CF (see middle panels in Supplementary Fig. 8). This made the 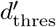 to be set to 3. The probe was never detected assuming only IHC dysfunction (the TMCs were arbitrarily set to the lowest masker level of 10 dB SPL; see Supplementary Fig. 9). All in all, the AN simulations cannot account for the behavioral data.

## Discussion

### Main findings and assumptions

The simulated TMCs and corresponding BM I/O functions in the present study did not align with behaviorally measured TMCs and corresponding BM I/O functions from Jepsen and Dau (2011). Simulated results were obtained using a state-of-the-art model of the AN (Bruce et al. 2018), whereas the behavioral results involved processing of the entire auditory system. It was considered that, in order for the behaviorally measured TMCs to show BM compression, enough information had to be present at the level of the AN, as the AN is a mandatory stage that forms an information bottleneck following the BM. In the simulations, we assumed that 1) information at the level of the AN was present exclusively in the form of spike rate and synchrony, 2) information was integrated across the whole population of the AN, and 3) all types of AN fibers, independent of their SR, could contribute to the final response. It was also assumed that, in order to detect the probe, the listener may compare AN activity evoked by the probe at a time window centered at the probe time versus the same corresponding time window in the masker-only block when the probe was not presented (green and blue rectangles in Fig. 1(b)). An alternative analysis was also performed comparing the probe time window against SR (see Supplementary Fig. 10). Also in this case, simulated TMCs did not align with the behavioral data, showing even more linear BM I/O functions. Other strategies than the ones analyzed, such as detection of discontinuity (or significant reduction) of rate or synchrony between the masker offset and the probe onset, cannot be discarded. Considering, however, the AN model to be a good approximation of the biological system, and accepting the assumptions indicated previously, the simulated results suggested that the compression estimated based on TMCs cannot be exclusively attributed to BM compression.

### Masker rate-level functions and post-masker recovery time courses

In the psychoacoustical domain, the main rationale of forward-masking based paradigms used to estimate cochlear compression relies on the assumption that the forward masking effect decays over time. The initial strength of such forward masking effect decay is dependent on the magnitude of the masker-induced BM vibration at the tonotopic location of the probe (on-CF). It is further assumed that a certain strength of the masking effect is required to mask the activity evoked by the probe. Because the forward masking effect decays over time, a higher masker level will be required with increasing MPI. For an off-CF masker, the growth at the tonotopic location of the probe is assumed to be linear with masker level. For an on-CF masker, it is assumed that the activity evoked by the masker grows compressively. Hence, a larger increment of masker level is required for the on-CF masker compared to the off-CF masker to increase the masking effect by the same amount (Nelson et al. 2001; Oxenham and Bacon 2003). Consistent with this, the behavioral TMCs for NH listener showed different slopes for the on- and off-CF maskers (Fig. 3 (a)).

In the physiological domain, the masker produces a reduction of neuronal activity at the masker offset (Smith 1977; Harris and Dallos 1979). This reduction does not only reduce the spontaneous activity of the AN, but also the activity evoked by the probe. The amount of reduction is dependent on the masker level and MPI conditions. Harris and Dallos (1979) systematically studied forward masking in single AN fibers in the chinchilla. They demonstrated that both the time course and the magnitude of the effect of forward masking (post-masker recovery) were associated to the spike rate evoked by the masker and not to its absolute physical magnitude. More crucial to the present study, they demonstrated that the time course and the magnitude of the post-masker recovery was independent of the spectral content of the masker (same for on- and off-CF maskers). It is unclear though whether their conclusions are valid for all AN fiber types or only for HSR fibers. Moreover, one could question whether the results acquired in single AN fibers can be directly extrapolated to the AN population response.

Figure 4 shows the analysis of the post-masker recovery time course of simulated HSR AN neurons tuned to on-CF (4 kHz). Panels (a) and (b) show rate as a function of time and masker level for the on-CF and the off-CF maskers, respectively. The results are shown as a surface plot as well as a contour plot with iso-rate slices. Panel (c) shows the same contour plots but shifting the off-CF response along the masker level axis to match the rate thresholds of the maskers. The time course of the post-masker recovery of rate showed similar profiles as a function of masker level for on- and off-CF masker, consistent with Harris and Dallos (1979). Panels (a) and (c) show that the rate evoked by the probe (at around 1.3 seconds in this example for a MPI of 100 ms) reduced as masker level increased, reaching eventually 0 spk/s. For MSR AN fibers, the post-masker recovery profiles were found also to be similar for on- and off-CF maskers (Supplementary Fig. 11). For LSR AN fibers, a clear post-masker recovery profile could not be observed due to the low SR of the fiber (Supplementary Fig. 12). The weak rate evoked by the probe in LSR fibers seemed to be still present at masker levels well above the on-CF masker threshold (panel (a)), whereas for the off-CF masker the rate induced by the probe seemed to be completely diminished as soon as the rate evoked by the off-CF masker was higher than SR. This could be explained by different growths of rate for on- and off-CF maskers due to BM compression. In the experimental paradigm simulated in the present study, the low physical intensity of the probe barely excited the LSR fibers (evoking a rate very close to SR), which led to very small *d*′ not contributing significantly to the final 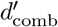 and to the simulated TMCs. The post-masker recovery time course depends directly on the rate evoked by the masker (Harris and Dallos 1979). Thus, the rate-level functions of the maskers can directly be linked to the effect of reduction of the rate evoked by the probe. Hence, the effect of on- and off-CF maskers on the rate-level functions of AN neurons tuned to the on-CF location needs to be different (with a shallower slope for the on-CF masker than for the off-CF masker) in order to reflect BM compression. In terms of the psychoacoustical interpretation, this translates to that the strength of the forward masking effect will increase “slower” with masker level for an on-CF masker than for an off-CF maskers (e.g., Oxenham and Bacon 2003; Nelson et al. 2001).

Figure 5 shows simulated rate-level functions of AN neurons tuned at the on-CF location for the on- and off-CF maskers. Panels (a-b), (c-d) and (e-f) show results for HSR, MSR and LSR AN fibers, respectively. Blue circles show responses to the on-CF masker and red squares to the off-CF masker, and corresponding solid lines indicate fits to the rate-level curves up to a stimulus level of 100 dB, as described in Yates et al. (1990). The rightmost panels (b, d, f) show overlapping rate-level functions by shifting the off-CF function so that both functions were equal at a rate of 30 spk/s above threshold. The overlapped rate-level functions showed identical growths for the HSR fibers (panel (b)) and very similar growth (except for the very high intensities) for the MSR fibers (panel (d)). In agreement with the post-masker recovery time course analysis shown in Fig. 4, on-CF BM compression was not reflected in the rate-level functions of HSR and MSR fibers in the current paradigm with a low-level probe and an on- vs off-CF masker frequency ratio of 0.6. The LSR rate-level functions (panel (f)) exhibited different slopes, with a shallower growth for the on-CF than for the off-CF masker, reflecting BM compression. These simulations agree with physiological data from AN neurons in the guinea pig (Yates et al. 1990). It was demonstrated that the narrow dynamic range of HSR fibers covers only the linear growth of the BM near threshold, and thus they do not reflect the BM compressive growth. This leads to identical rate-level functions for on- and the off-CF maskers. MSR and LSR fibers can reflect the compressive growth of the BM, but the difference between rate-level functions for on- and off-CF maskers depends on the frequency ratio between them. Supplementary Fig. 16 shows a qualitative good correspondence between the simulated rate-level functions using the cat version of the AN model and the physiological results in guinea pigs reported in Yates et al. (1990), besides inter-species differences. The simulated TMCs (Fig. 3) obtained after combining all types of AN fibers indicated that the HSR fibers were the main contributor to the final TMC due to the fixed low level of the probe. This also resulted in a no contribution of LSR fibers to the final simulated TMCs (see also Supplementary Fig. 12). Rate turned out to be the most contributing metric, as synchrony may be an unreliable metric due to the short duration of the probe. Thus, the close to parallel growth observed in the simulated on- and off-CF TMCs could be explained by the compressive growth of the BM not being reflected in the rate of the HSR and MSR fibers in the current paradigm.

### On the discrepancy between psychoacoustics and AN physiology

A potential explanation for the mismatch between the psychoacoustical rationale and the findings from AN physiology and the current modeling study could be a discrepancy in the mechanism of masking. In psychoacoustics, tone-in-noise detection has been often considered to follow a model similar to the power spectrum model (Fletcher 1940). In this model, the auditory system can detect a signal if the energy of the signal exceeds the energy of the noise (i.e., a certain signal-to-noise ratio (SNR) is achieved) in one auditory filter. Applied to the current forward masking paradigm, the power spectrum model would assume that both, the probe and the masker, excite the auditory filter centered at the probe frequency (on-CF). The energy of the masker would decay after the masker offset to account for the behavioral exponential decay of forward masking (Oxenham and Moore 1994; Nelson et al. 2001; Lopez-Poveda et al. 2003). This concept has successfully been used to model psychoacoustical forward masking by combining an on-CF peripheral nonlinearity, such as a time-invariant power law (Oxenham and Moore 1994) or a compressive nonlinearity (Oxenham et al. 1997), with a linear temporal integration window representing a more central processing (Oxenham and Moore 1994). In this model, the peripheral nonlinearity scales the energy of the masker differently for on- and off-CF maskers, and the temporal integration window results in a decayed masker energy towards the probe time in the auditory filter tuned to on-CF. When the decayed masker energy is above a certain value relative to the constant energy of the probe, the probe will be masked.

This is where the psychoacoustical concepts and the physiological findings deviate. While in the psychoacoustical concept it is assumed that the activity evoked in the auditory filter centered at the probe frequency is constant, and the overall SNR depends on the masker level and the MPI, in physiology it is the activity evoked by the probe that is reduced by the masker-offset reduction in rate.

The low sound level of the probe excited the HSR and MSR fibers, but neither of them were affected by the compressive nonlinearity of the BM (Fig. 5) (see also Supplementary Fig. 16 and Harris and Dallos 1979). MSR fibers could reflect the BM compressive nonlinearity but, according to the model simulation, not with the 0.6 masker frequency ratio used in the TMC paradigm in Jepsen and Dau (2011). In addition, some physiologists have questioned whether MSR and LSR AN fibers play a significant role in sound encoding, as their rate-level functions do not grow steep enough to encode intensity changes and most of their upward projections do not belong to the ascending pathway (Carney 2018).

Assuming that the simulated results presented here are a realistic representation of the post-masker reduction in rate in the human AN, adding a temporal integration window will not lead to a change in the slopes of the simulated TMCs (Fig. 3), as the temporal integration window is a linear processing. The idea of combining a physiological model of the auditory periphery with the temporal integration window model was shown to be successful in simulating TMCs for NH and HI listeners (Jennings et al. 2014). However, interestingly, Jennings et al. (2014) did not fed the output of the AN to the temporal integration window but the output of the IHC. At the output of the IHC (which provides the IHC transmembrane potential) the voltage evoked by the probe was constant and unaffected by the masker level (consistent with the psychoacoustical concept). Hence, it could be used as a reference excitation from which to compare the growth of the maskers (see Supplementary Fig. 13). This approach circumvents the effects found at the level of the AN that the probe evoked neuronal activity reduces with increasing masker level (Fig. 4). Also, the IHC output had an unlimited dynamic range such that the growth of both the on- and off-CF maskers was continuous across all masker levels and that the compressive nonlinearity of the BM could be reflected (Supplementary Fig. 13). This was not the case in the AN, where the different types of neurons show different thresholds and limited dynamic ranges (Liberman 1978).

Considering that, as our modeling analysis suggested, TMCs may grow close to parallel at the level of the AN, an obvious question arises. What explains the consistently measured behavioral TMCs (e.g., Nelson et al. 2001; Lopez- Poveda et al. 2003; Plack et al. 2004; Lopez-Poveda et al. 2005; Lopez-Poveda and Alves-Pinto 2008; Jepsen and Dau 2011; Lopez-Poveda and Johannesen 2012; Johannesen et al. 2014; Fereczkowski et al. 2017)? Answering this question is clearly beyond the scope of the current study. However, one could speculate that a central nonlinear process could lead to similar net results. In the behavioral model of forward masking (Oxenham and Moore 1994), the peripheral compressive nonlinearity was coupled to a central linear process. Our modeling analysis suggested that, even though there was a compressive nonlinearity in the cochlea, the responding neurons in the AN were not affected by it, resulting in a *de facto* close to linear process. This linked to a central nonlinearity could theoretically lead to the same overall result in the percept, that is, at the end of auditory pathway. Results from non-human mammals have shown that the effect of forward masking in the AN did not correspond to the effect observed behaviorally (Relkin and Turner 1988). In contrast, the addition of more central processes, which may include inhibitory mechanisms at the level of the IC, resulted in strikingly similar forward masking effects to the effects found perceptually (Nelson et al. 2009). Potentially, projections from offset-type inhibitory neurons from the superior paraolivary nucleus (SPON) to IC neurons could further reduce the probe evoked response observed at the level of the AN. These type of offset neurons were shown to be sharply frequency tuned at threshold and to become more broadly tuned at higher stimulus levels, which could lead to frequency dependent effects (Kulesza et al. 2003). If this was to be the case, it would imply that the compression estimated behaviorally using the TMC paradigm would not reflect the BM compressive nonlinearity, but rather inhibitory mechanisms at the level of the IC. Interestingly, such inhibition at the IC level was shown to be diminished after acoustic trauma resulting in hearing impairment (Wang et al. 2002). A similar modeling analysis as presented here including neurons from more central stages (cochlear nucleus (CN), IC, SPON) (Salimi et al. 2017) could shed light on the underlying mechanisms of TMCs and forward masking and their potential use to estimate BM compression.

## Conclusions

- Based on the stated assumptions and the results of the present study we concluded that:
- Psychoacoustical TMCs could not be simulated using a state-of-the-art model of the AN using spike rate and synchrony as decision metrics.
- A low-level probe in a TMC paradigm excites the HSR and some MSR fibers, but not the LSR fibers. Animal studies suggest that HSR fibers do not reflect BM compression. Our modeling results also supported this fact. Furthermore, the modeling simulations suggested that MSR fibers with a frequency ratio of 0.6, as used in the TMC paradigm, do not reflect BM compression either. As a result, the simulated TMCs grew close to parallel. Hence, it seems reasonable to assume that TMCs using the current paradigm do not reflect cochlear compression.
- The numerical match in BM compression and behaviorally estimated compression using TMCs suggested that may exists another source of compression which may contribute to the compression measured in the behavioral tasks.
- The results of the present study and the behavioral paradigms might help to disentangle peripheral compression (BM and AN) and more central (IC) sources of compression.

## Data Availability

Simulated results can be facilitated to any interested researcher on demand.

## Acknowledgements

This work was funded by the Novo Nordisk Foundation grant NNF17OC0027872.

## Supplementary Material

**Figure 6.**
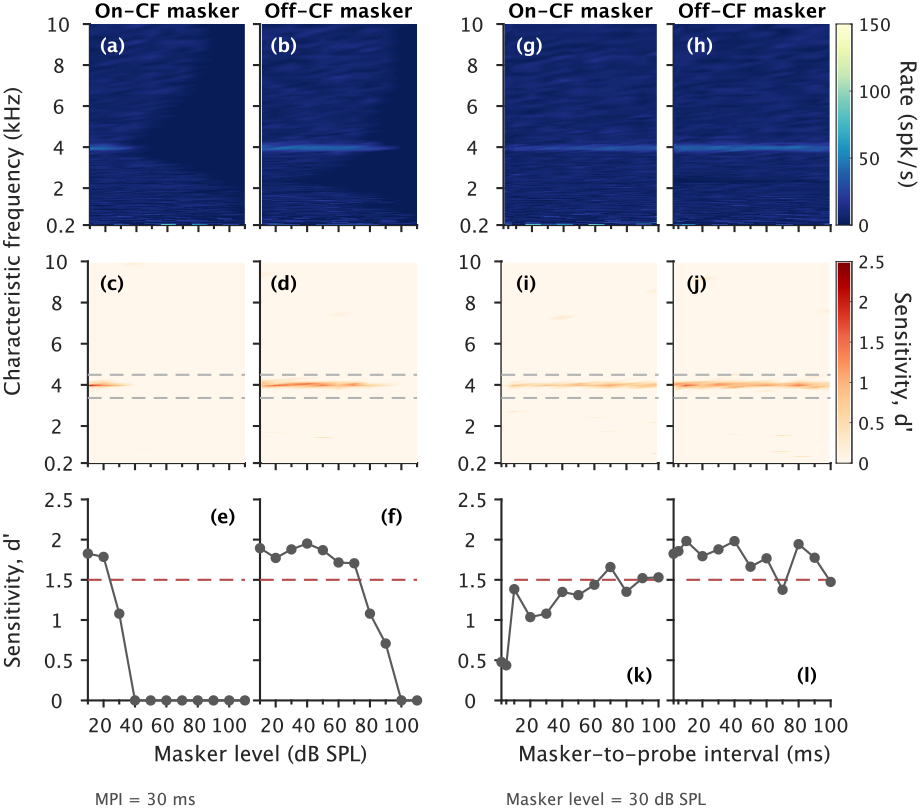
Same as Fig. 2 but for MSR AN fibers.

**Figure 7.**
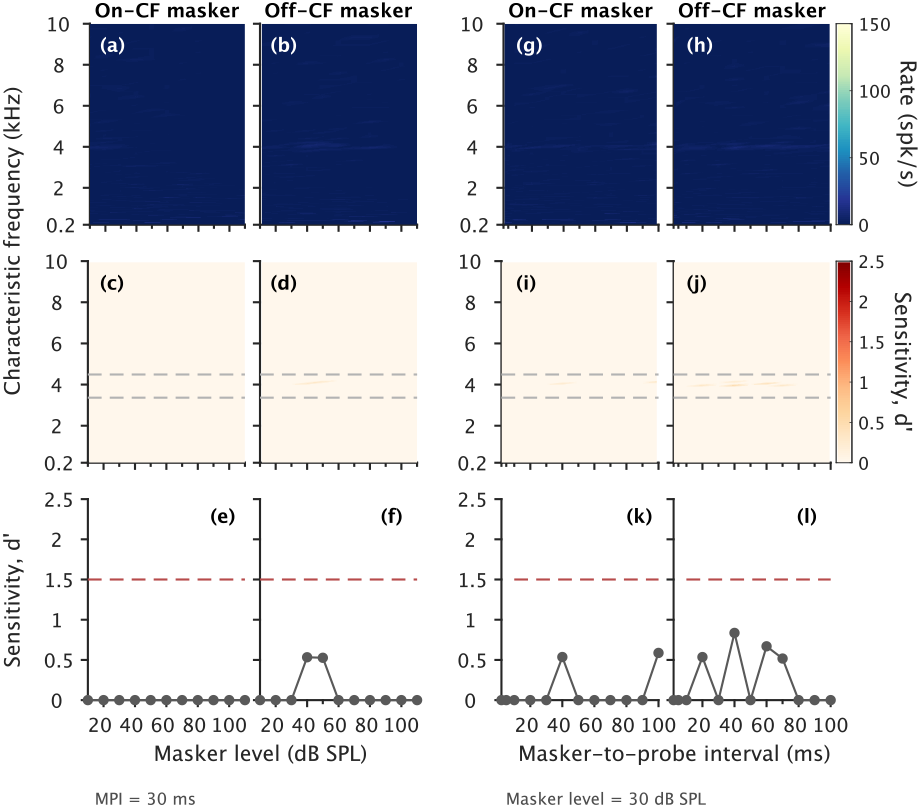
Same as Fig. 2 but for LSR AN fibers.

**Figure 8.**
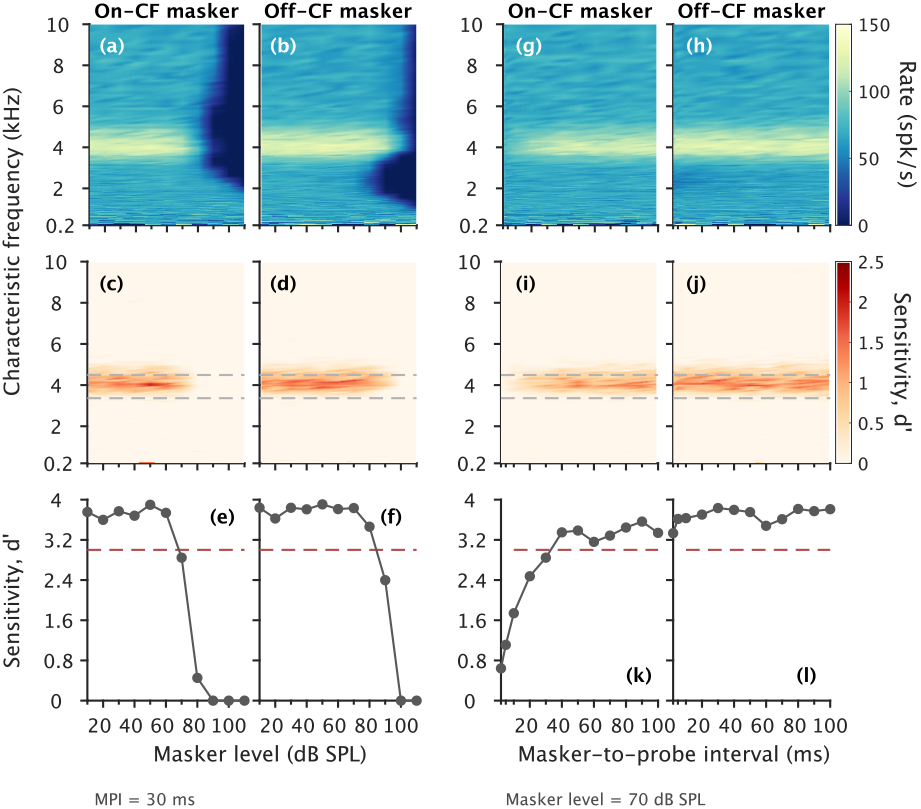
Same as Fig. 2 but for a HI listener accounting for the hearing threshold elevation assuming only OHC dysfunction.

**Figure 9.**
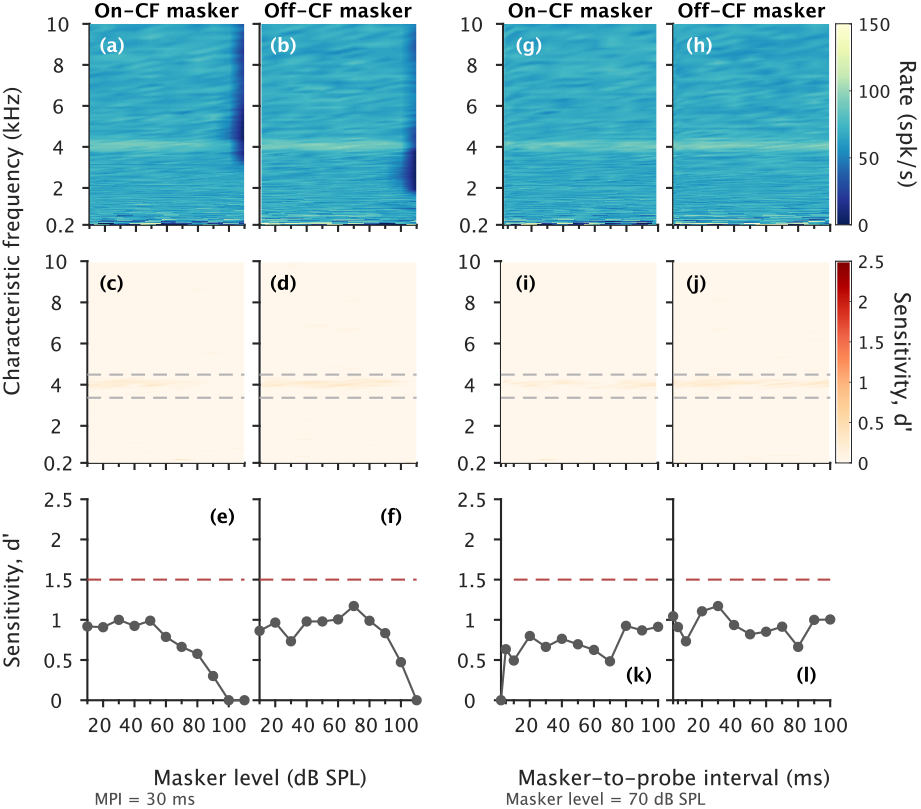
Same as Fig. 2 but for a HI listener accounting for the hearing threshold elevation assuming only IHC dysfunction.

**Figure 10.**
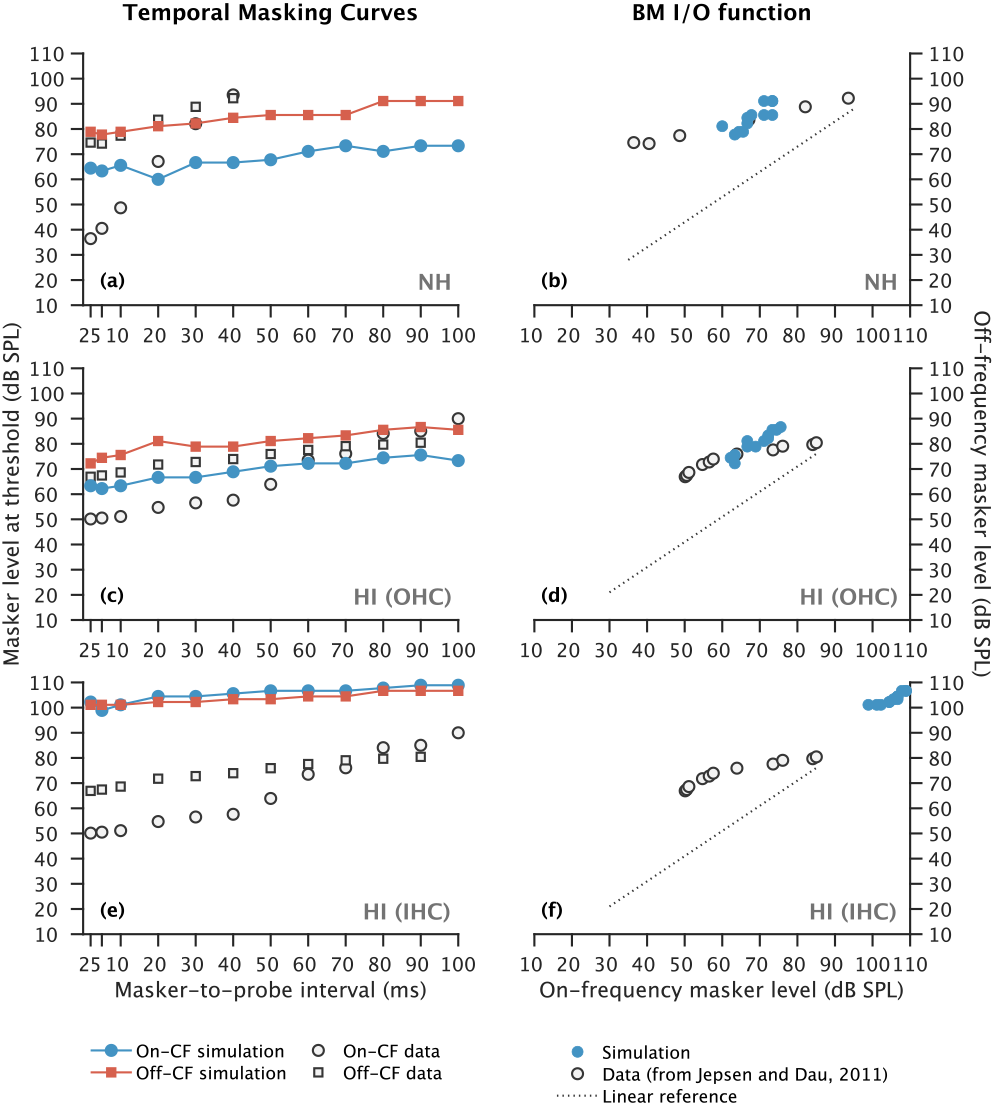
Same as Fig. 3 but using a window at SR (yellow rectangle in Fig. 1(b)) instead of the corresponding probe window in the masker-only block (blue rectangle in Fig. 1(b)).

**Figure 11.**
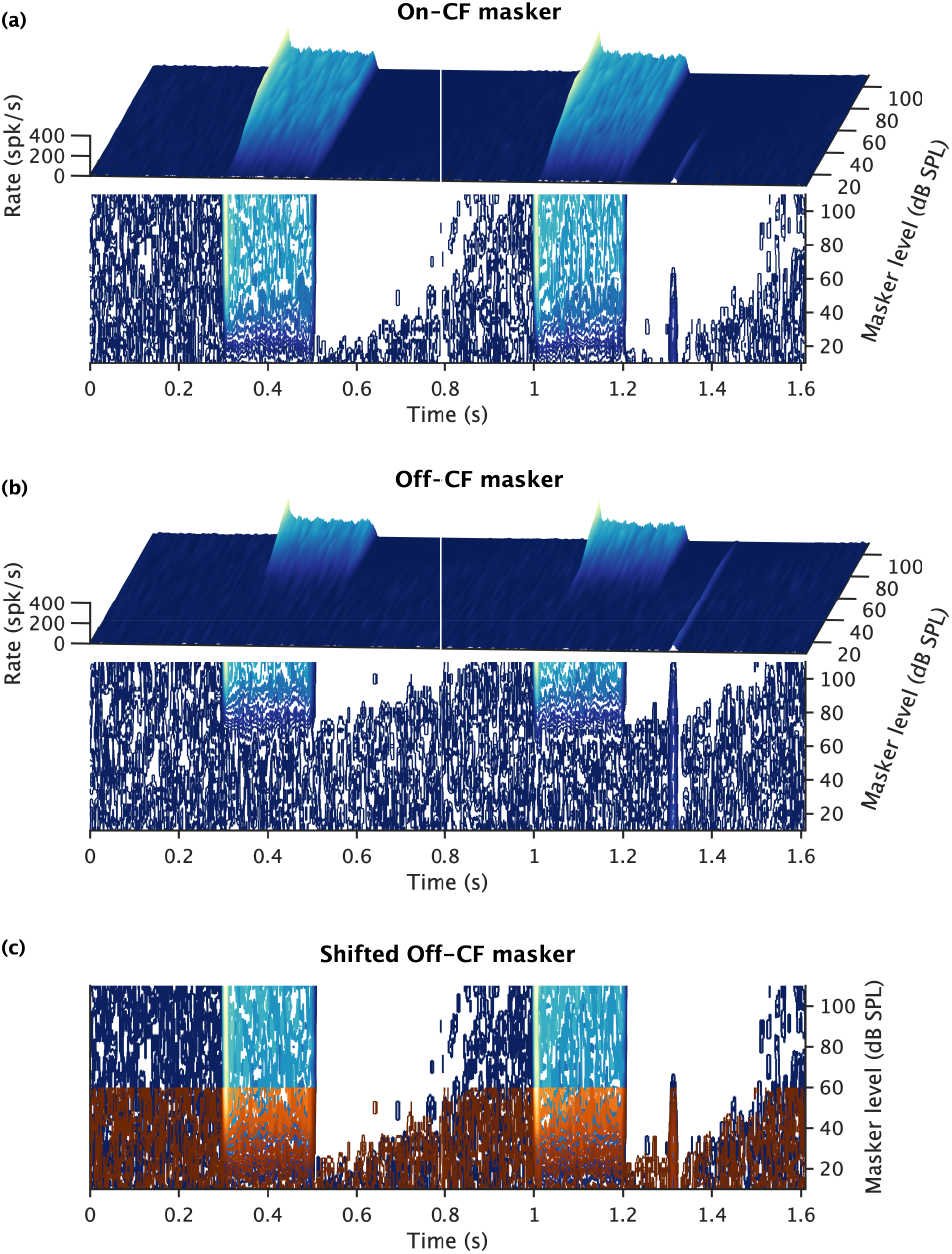
Same as Fig. 4 in the main text but for MSR fibers.

**Figure 12.**
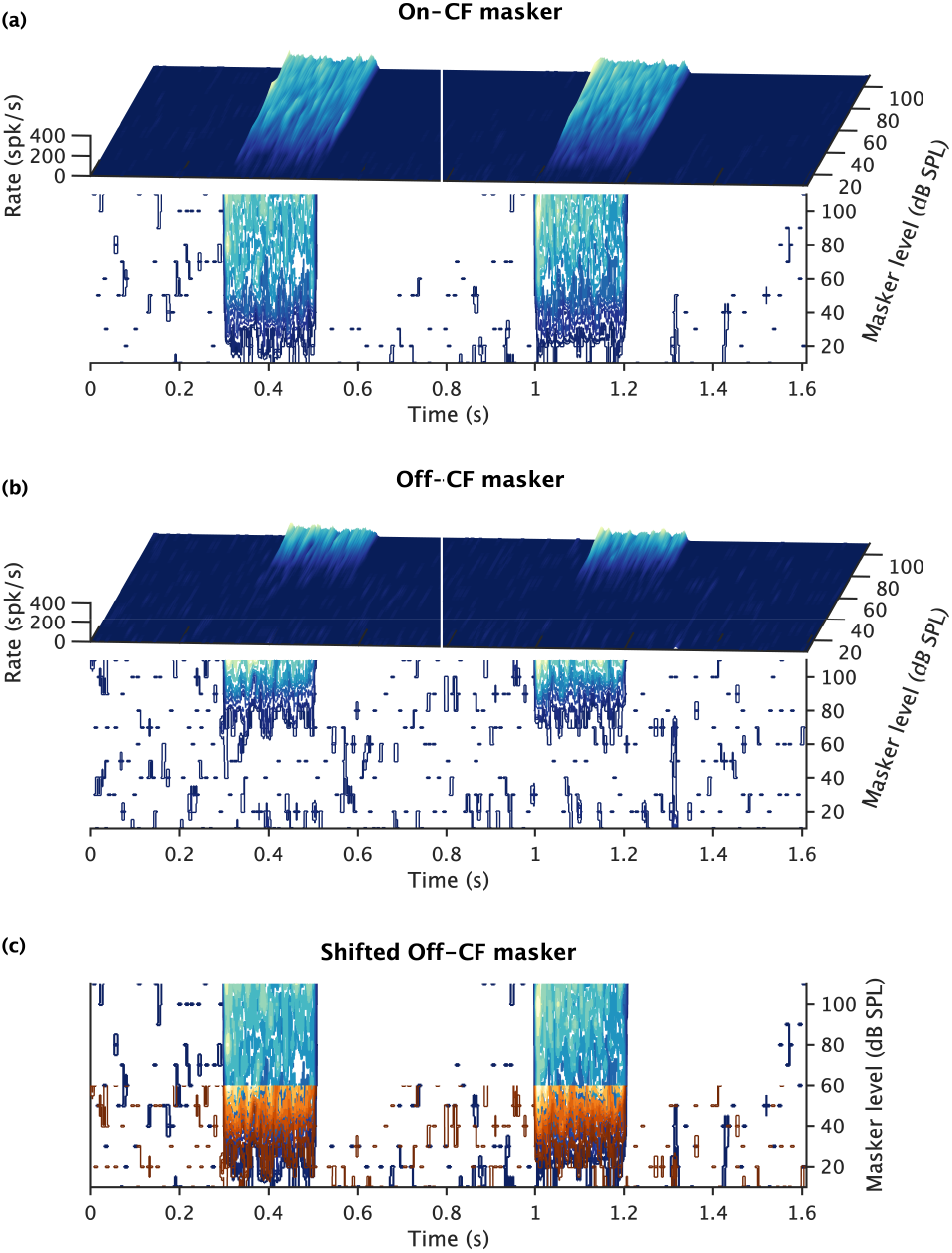
Same as Fig. 4 in the main text but for LSR fibers.

**Figure 13.**
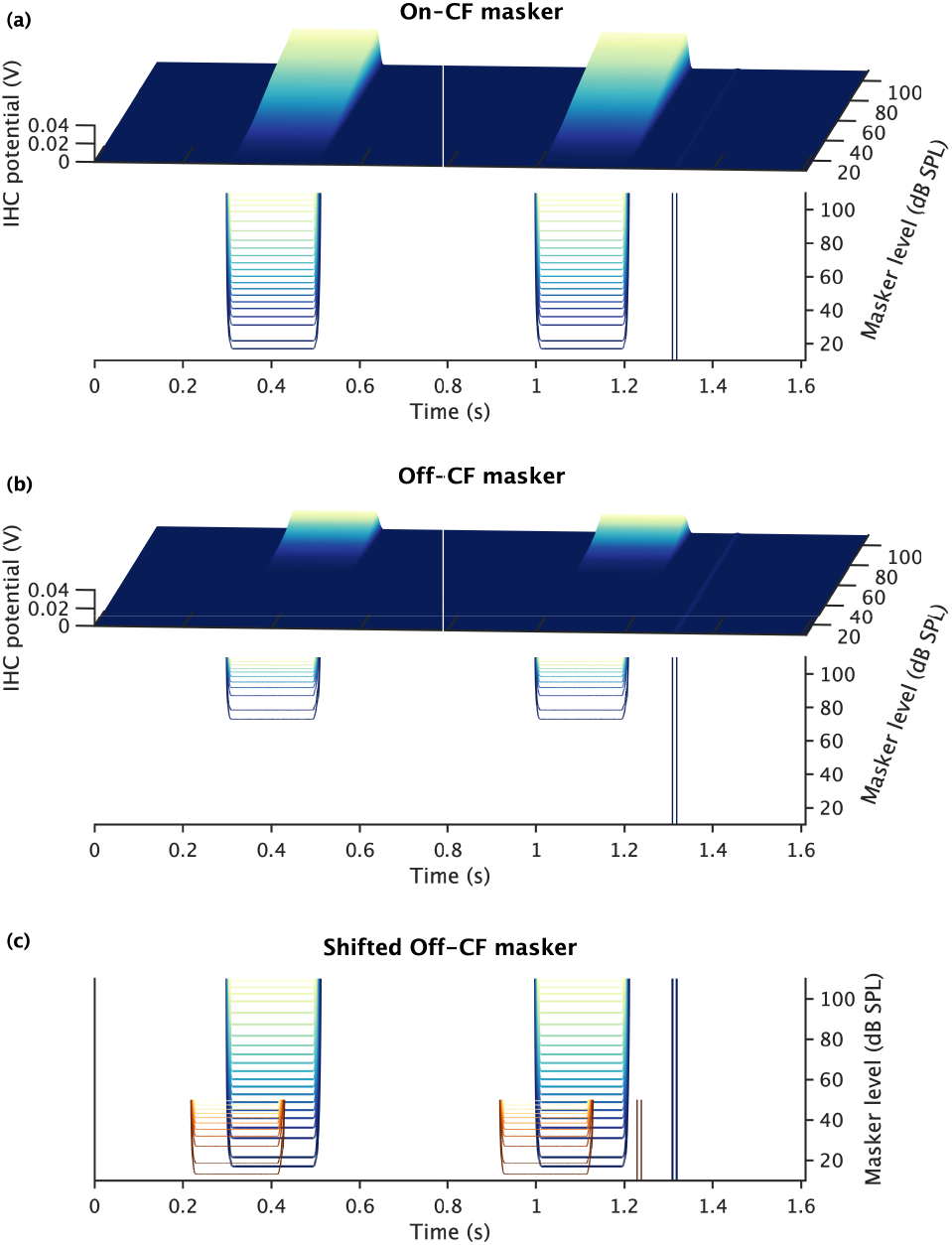
Same as Fig. 4 in the main text but for the model output simulating the IHC transmembrane potential. The off-CF contour plot in panel (c) was advanced 80 ms in time to help on visualizing the traces.

**Figure 14.**
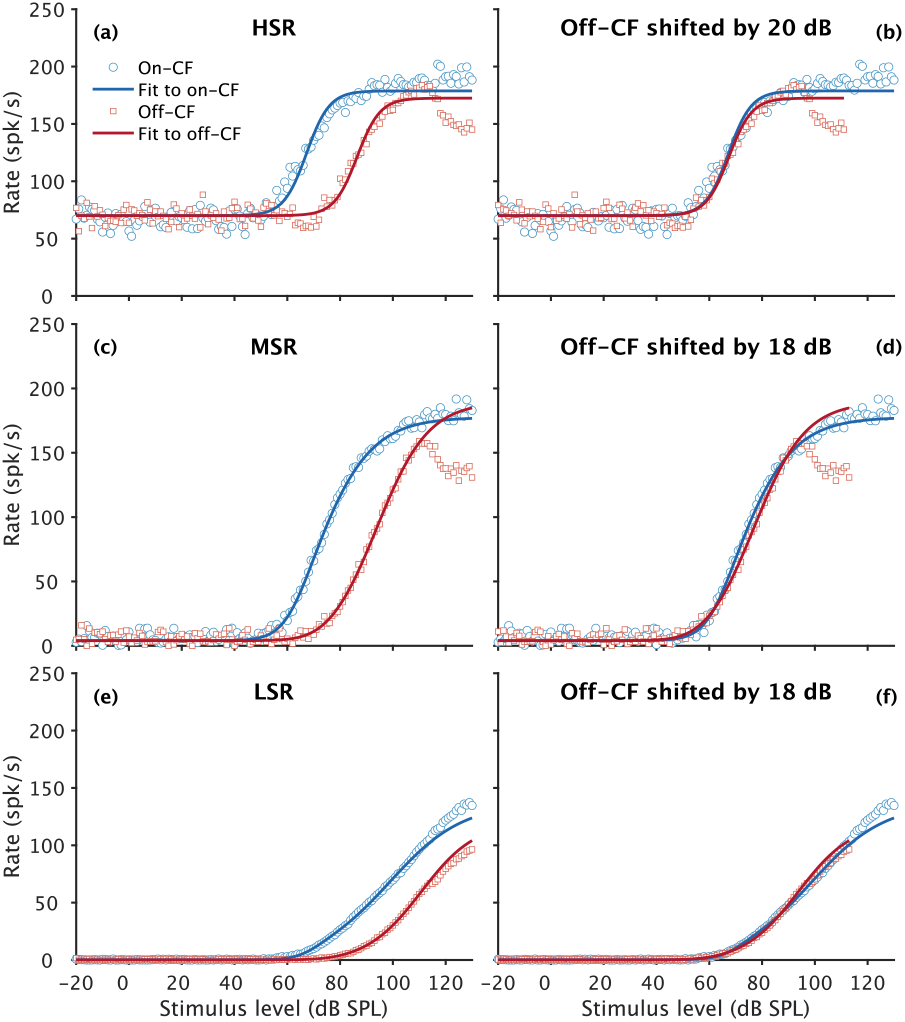
Same as Fig. 5 in the main text but for the HI listener assuming that the hearing threshold elevation is due to OHC dysfunction only.

**Figure 15.**
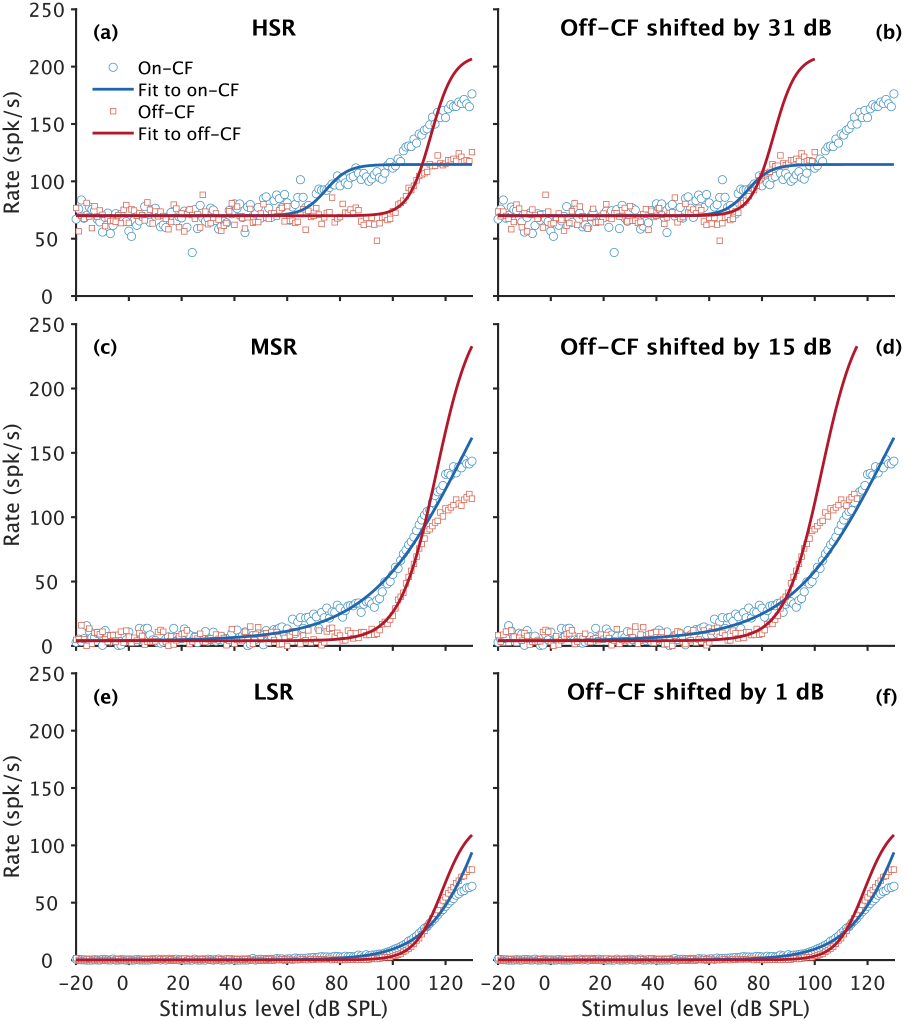
Same as Fig. 5 in the main text but for the HI listener assuming that the hearing threshold elevation is due to IHC dysfunction only.

**Figure 16.**
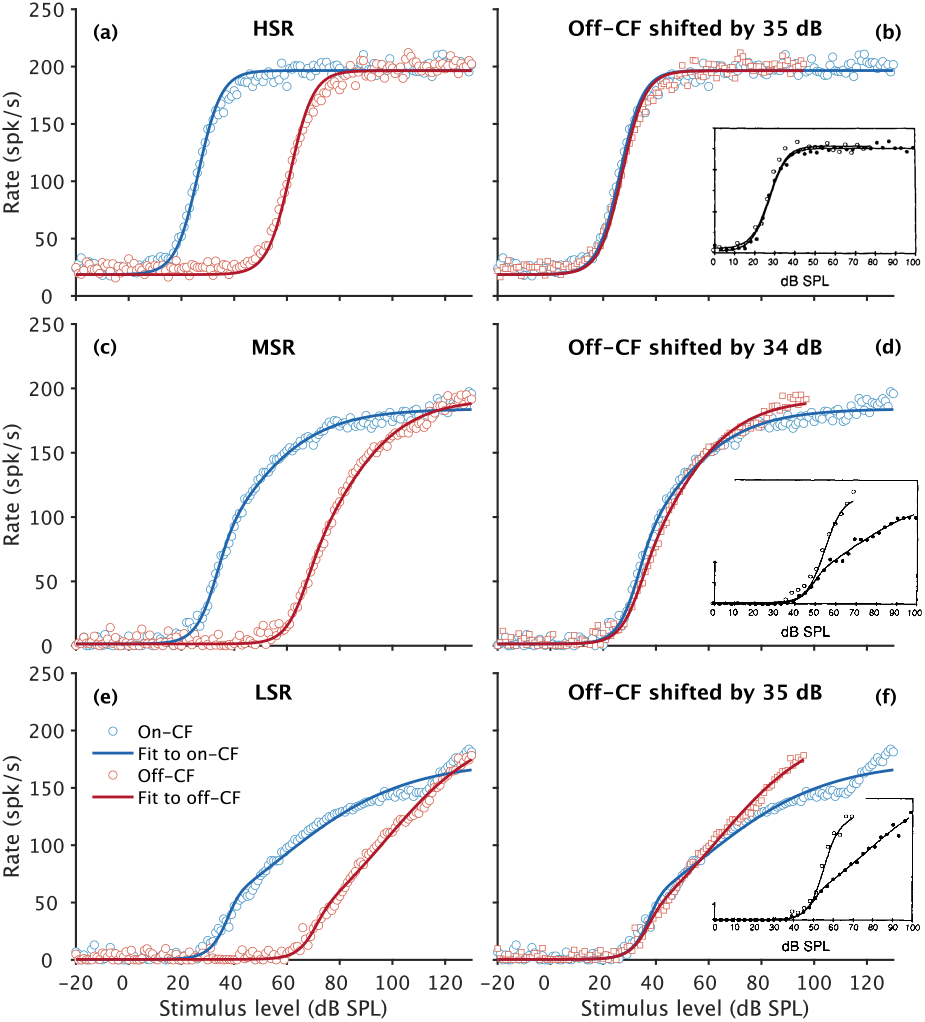
Same as Fig. 5 but using the stimulus as in Fig. 1 in Yates et al. (1990). The cat version of the AN model was used to simulate the rate-level functions recorded in guinea pigs. The stimulation frequencies used in the recordings (guinea pig: on-CF of 20 kHz and off-CF of 17 kHz) were transformed into the cat by selecting the the same relative cochlear location from the base based on Greenwood (1990) (cat: on-CF of 26.6 kHz and off-CF of 22.6 kHz). These frequencies result in an off-CF on-CF frequency ratio of 0.85. Insets show the original data in guinea pigs from Yates et al. (1990) (reprinted with permission from the authors and the publisher).

